# Long COVID symptoms from Reddit: Characterizing post-COVID syndrome from patient reports

**DOI:** 10.1101/2021.06.15.21259004

**Authors:** Abeed Sarker, Yao Ge

## Abstract

**Objective:** To mine Reddit to discover long-COVID symptoms self-reported by users, compare symptom distributions across studies, and create a symptom lexicon.

**Materials and Methods:** We retrieved posts from the */r/covidlonghaulers* subreddit and extracted symptoms via approximate matching using an expanded meta-lexicon. We mapped the extracted symptoms to standard concept IDs, compared their distributions with those reported in recent literature and analyzed their distributions over time.

**Results:** From 42,995 posts by 4249 users, we identified 1744 users who expressed at least 1 symptom. The most frequently reported long-COVID symptoms were *mental health-related symptoms* (55.2%), *fatigue* (51.2%), *general ache/pain* (48.4%), *brain fog/confusion* (32.8%) and *dyspnea* (28.9%) amongst users reporting at least 1 symptom. Comparison with recent literature revealed a large variance in reported symptoms across studies. Temporal analysis showed several persistent symptoms up to 15 months after infection.

**Conclusion:** The spectrum of symptoms identified from Reddit may provide early insights about long-COVID.

## INTRODUCTION

Many patients continue to experience symptoms long after the acute phase of infection with the SARS-CoV-2 (COVID-19) virus,^1,2^ and currently, the persistence of symptoms 28 days after the diagnosis of COVID-19 infection is referred to as ‘*post-acute COVID syndrome*’, ‘*post-acute sequelae of SARS-CoV-2*’, ‘*long-haul COVID-19*’ or ‘*long-COVID*’.^3^ A growing body of research is exploring long-COVID and the constellation of symptoms that patients experience following the acute phase of COVID-19 infection.^3–6^ The phrase ‘long-COVID’ was coined by patients, many of whom initially suffered mild symptoms only to progress to complex illnesses.^7^ To date, research suggests that long-COVID symptoms may affect people of varying ages including children and those with asymptomatic acute infections.^8–12^ However, after more than a year since the beginning of the pandemic, there are still many unknowns associated with long-COVID, including the full spectrum of symptoms that patients experience, their progression, and their long-term manifestation. As more patients around the world continue to recover from acute COVID-19 infections, long-COVID is emerging to be a complex health problem affecting millions of people globally, and a wide range of symptoms are being discovered over time.^5,9,12,13^ Consequently, this syndrome is emerging as a critical research topic relevant to the pandemic with many ongoing studies globally.

One potential source of information regarding long-COVID syndrome is social media, as many people, including healthcare professionals who had been infected with COVID-19, are reporting their experiences through this medium.^14,15^ Social media adoption is currently at an all-time high and continues to grow globally and in the United States.^16,17^ In the past, large volumes of user-generated information on social media have been utilized by researchers to obtain early insights about health-related topics, including infectious diseases.^18,19^ Due to the global nature of the current COVID-19 pandemic, there is an abundance of knowledge about diverse aspects of it in publicly available social media data. Studies have thus attempted to utilize this resource to answer targeted questions associated with COVID-19.^20–23^ In this study, we attempted to mine publicly available social media data from Reddit to discover and analyze the spectrum of symptoms self-reported by sufferers of long-COVID. This study builds on our prior work on the topic, which focused on extracting self-reported COVID-19 symptoms from Twitter.^23^ Our objectives for this study were to extend a COVID-19 symptom lexicon built using Twitter data, deploy the extended lexicon to identify self-reported long-COVID symptoms from a specific forum on Reddit, analyze the distribution of symptoms and compare them with symptoms reported in recent studies. We describe our methods and findings in the following sections.

## MATERIALS AND METHODS

### Data source and collection

We collected data for study from Reddit, which consists of many topic-specific forums called subreddits. The subreddit */r/covidlonghaulers* has emerged as the go-to forum for the discussion of long-COVID related topics, and it is an information-rich source for obtaining self-reported long-COVID symptoms. As of 6^th^ June, 2021, the subreddit had over 14,000 subscribers and over 1200 threads, most of which focus on long-COVID experiences self-reported by users. Within each thread, users report and discuss the symptoms they experience and also often provide timelines of their symptoms. This subreddit is thus an excellent resource for obtaining crowdsourced information about long-COVID syndrome. We collected all the posts from this subreddit using the PRAW application programming interface (API).^24^ The earliest post included in our analyses was from 25^th^ July, 2020 and the latest post was from 6^th^ June, 2021.

### Symptom extraction from user posts

We first manually annotated a symptom lexicon from Reddit, grouping similar symptoms and mapping each symptom expression to a standard identifier in the unified medical language system (UMLS) using the National Center for Biomedical Ontology BioPortal.^25^ We augmented the lexicon entries for rarely occurring symptoms by adding expressions from the Medical Dictionary for Regulatory Activities (MedDRA).^26^ We combined this extended Reddit lexicon with the previously-developed Twitter lexicon^23^ to create a meta-lexicon consisting of common COVID-19 and long-COVID symptom expressions. We then further expanded the meta-lexicon automatically using the LexExp tool.^27^ During the manual annotation process, we discovered that users often expressed the resolution or absence of symptoms using negation expressions such as ‘*no*’, ‘*do not*’, ‘*never had*’, and so we created a small lexicon of such negations. To detect symptoms from free text, we applied an inexact matching method. Exact matching on social media free texts typically results in low recall due to the presence of nonstandard expressions and misspellings. To overcome this issue, in line with our past work, we searched through term sequence windows in the texts and computed the similarity of each text window with all entries in the meta-lexicon. Text windows that obtained similarities above a specific threshold with any entry in the lexicon were extracted and considered to be candidates for long-COVID symptoms. We used the Levenshtein ratio metric to compute similarity, with a high threshold of 0.98. For an expression of *n* terms, the term sequence windows were of the range [*n* − 1: *n* + 2]. A negation detection algorithm then searches for the occurrence of negations in the same post as the candidate symptom and estimating if the symptom appears within the scope of the negation. The negation detection algorithm preprocesses the texts by lowercasing and tokenizing, and labels any term appearing within a 3 token window or before an end-of-sentence marker following the negation term (eg., a period) as negated. We grouped the symptoms per user and generated their frequency distributions for analysis and comparison.

We evaluated the performance of the algorithm using the precision, recall and F_1_-score metrics. To estimate precision and recall, we selected a random sample of posts, including posts from which at least one symptom was automatically detected and posts from which our algorithm did not detect any. The F_1_-score is the harmonic mean of precision and recall (*F*_1_ − *score* = (2 × *precision* × *recall*)/(*precision* + *recall*)).

### Symptom distribution by month

During our exploration of the threads within the subreddit, we discovered many thread titles that specified the number of months since the initial COVID-19 diagnosis that the particular user was experiencing symptoms for (*eg*., ‘*8 month update*’, ‘*11 month long hauler just got first vaccine*’). We created a set of regular expressions to detect such expressions and computed the relative frequency distributions for the numbers of users expressing at least one symptom within these posts. We grouped the frequencies by month, starting from 3 up to 15.

## RESULTS

We collected 42,995 posts from 1220 threads and 4249 users. We reviewed posts associated with a total of 450 symptom expressions (*i*.*e*., the same post could be reviewed multiple times in association with different detected symptoms) along with 25 posts without any detected symptoms to estimate the performance of our approach (recall = 0.93, precision = 0.95, F_1_-score = 0.94). 1744 users expressed at least 1 non-negated symptom (2576 users without accounting for negations). Figure 1 presents the symptom distributions reported by this cohort. From the histogram (left) we can see that most users reported 1 – 6 symptoms. The median number of symptoms reported per user was 3 (boxplot; right) and a handful of users reported 12 or more symptoms, with 32 being the highest number of symptoms expressed by a single user.

**Figure 1.**
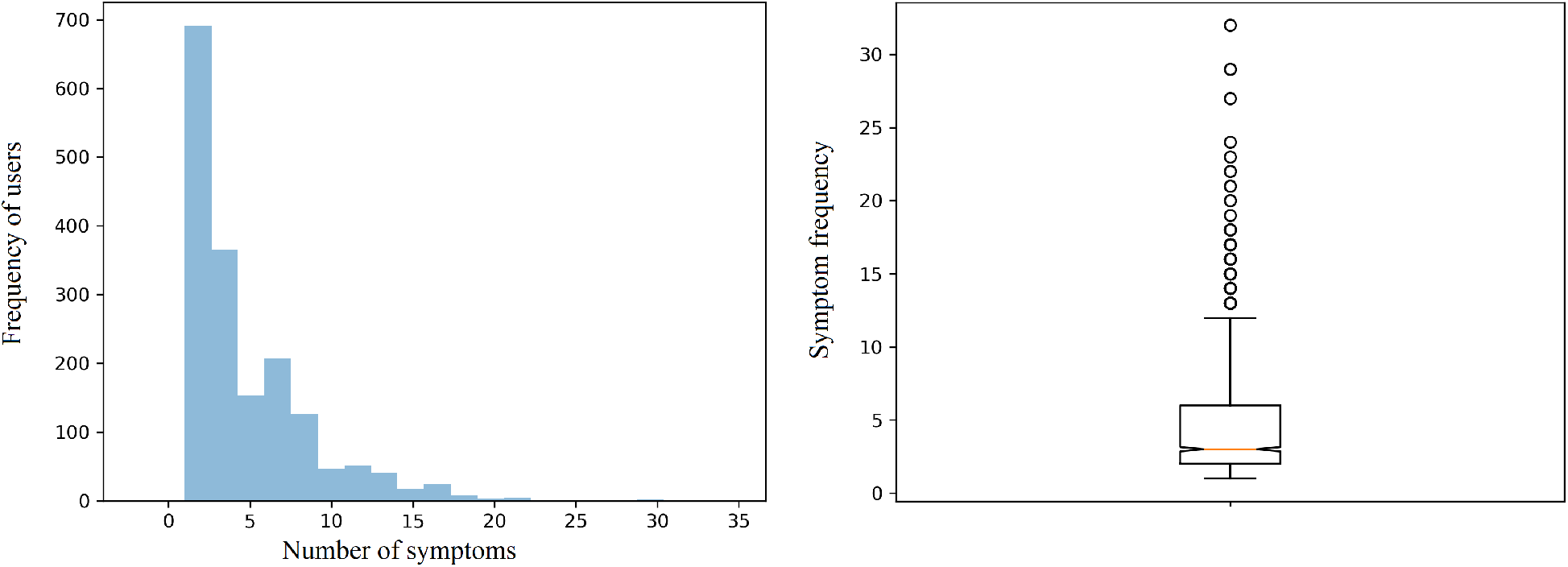
Distribution of long-COVID symptoms reported by Reddit users.

Figure 2 presents the raw counts for the top 20 symptoms reported by the users and their relative frequencies as a percentage of the total number of users who reported at least 1 non-negated symptom. There are some interesting differences between the acute COVID-19 symptoms reported on Twitter, which we presented in a prior publication, and the long-COVID symptoms we detected in this study.^23^ The most commonly reported long-COVID symptoms are *anxiety/stress and related mental health symptoms*.^†^ The next 4 most commonly reported symptoms are *fatigue, general & body ache & pain, confusion/disorientation*^‡^ and *dyspnea. Pyrexia*, which was the most commonly reported symptom among users who self-reported to have tested positive for COVID-19, is reported much less frequently for long-COVID (6^th^).

**Figure 2.**
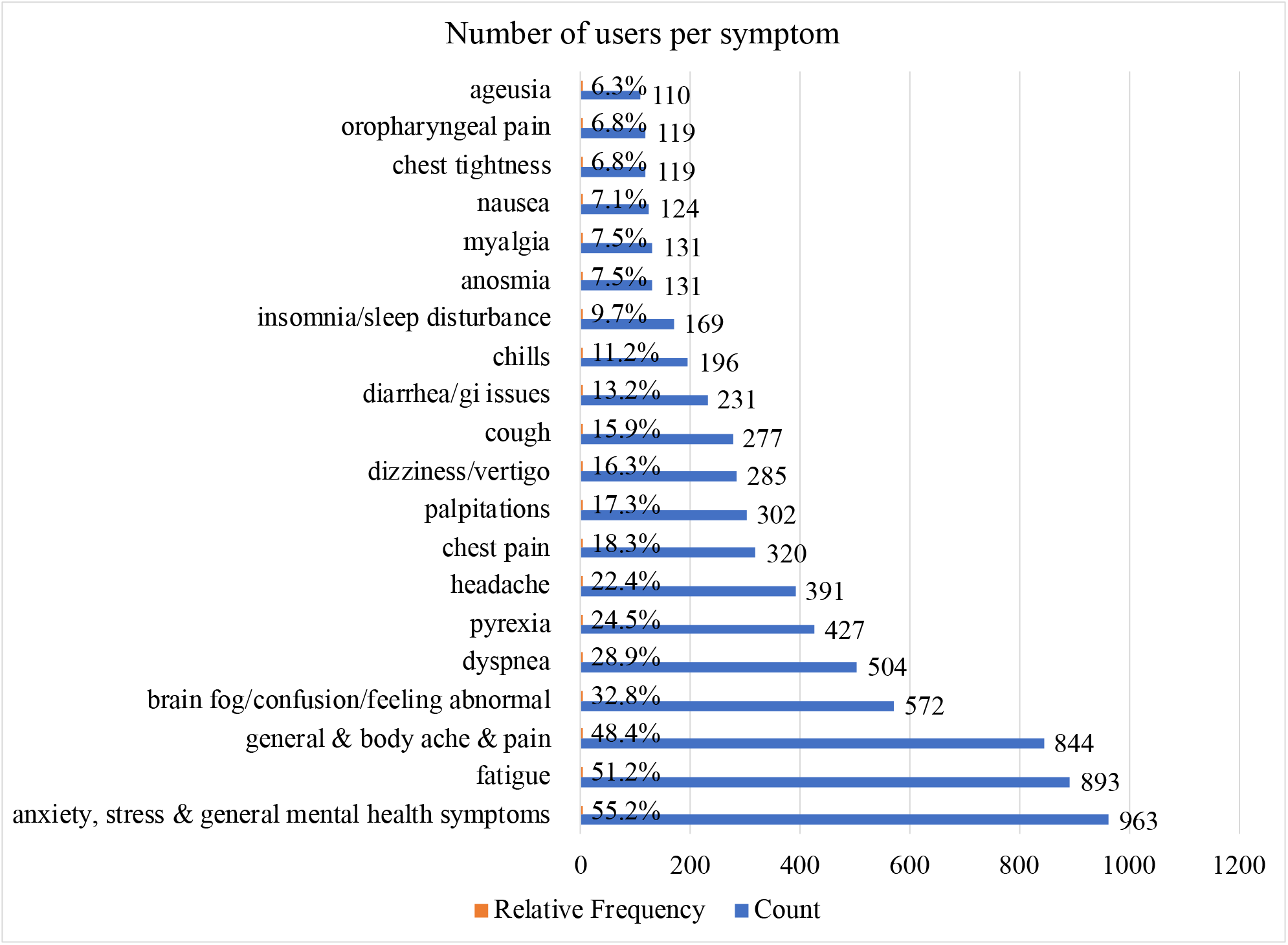
Number of users reporting each of the top 20 symptoms and their percentages.

Figure 3 illustrates, via a heatmap, the relative frequencies of 24 symptoms from months 3 to 15. From the figure, it appears that *general* & *body ache & pain, fatigue, anxiety, stress and other mental health issues*, and *dyspnea* were reported to be the most persistent symptoms, with a large number of users reporting to experience it 12 months after their initial diagnosis. Some symptoms, such as pyrexia, *GI issues*, and *oropharyngeal pain* appear to lower over time. Table 1 presents the relative frequency distributions of 10 commonly-reported long-COVID symptoms from this study and 7 recently-published papers on long-COVID. *Fatigue* is consistently reported with high frequency across studies, but considerable variations are can be observed in the distribution all symptoms.

**Table 1.** Comparison of long-COVID symptom distributions (in percentage) identified in this study with those reported in recent literature for frequently-reported symptoms.

|  | This study<br>(n=1744) | Osikomaiya et al.<sup>29</sup><br>(n=274) | Huang et al.<sup>30</sup><br>(n=1733) | Sykes et al.<sup>31</sup><br>(n=134) | Mandal et al.<sup>32</sup><br>(n=276) | Carfi et al.<sup>33</sup><br>(n=143) | Orrù et al.<sup>28</sup><br>(N=152) | Garrigues et al.<sup>34</sup><br>(n=120) |
| --- | --- | --- | --- | --- | --- | --- | --- | --- |
| Fatigue | 51.2 | 12.8 | 63 | 39.6 | 69 | 53.1 | 74.3 | 55.0 |
| Ache & pain | 48.4 | 8.8* | 9<sup>†</sup> | 51.5* | N/A | 27.3<sup>†</sup> | 61.2* | N/A<sup>✱✱</sup> |
| Brain fog/confusion or dizziness | 32.8 | 5.2** | 6<sup>††</sup> | 25.4** | N/A | 8<sup>***,✱</sup> | 48.7 | 26.7** |
| Dyspnea | 28.9 | 9.5 | N/A | 59.7<sup>§</sup> | 53<sup>§</sup> | 43.4 | 40.1 | 41.7 |
| Fever/pyrexia | 24.5 | 6.2 | <1 | 10.4 | N/A | N/A<br>(excluded from study) | 19.1 | N/A |
| Headache | 22.4 | 12.8 | 2 | N/A | N/A | 10<sup>***</sup> | 46.7 | N/A |
| Insomnia/sleep disturbance | 9.7 | 9.8 | 26 | 35.1 | N/A | N/A | N/A<sup>✱✱✱</sup> | 30.8 |
| Chest pain | 18.3 | 9.8 | 5 | 17.9 | N/A | 21.7 | 26.3 | 10.8 |
| Cough | 15.9 | 9.2 | N/A | 35.1 | 34 | 18<sup>***</sup> | 21.1 | 16.7 |
| Palpitations | 17.3 | 7.4 | 9 | N/A | N/A | N/A | 38.8 | N/A |

\* Reported symptom is *myalgia*
\*\* Reported as attention or memory deficit or disorder
† Only joint pain (*arthralgia*) reported
†† Only dizziness reported
§ Reported as breathlessness
\*\*\* Estimated from figure
✱ Reported as *vertigo*
✱✱ Patients reported pain/discomfort using a 5 point scale
✱✱✱ Sleep disturbances were measured based on ISI scores

**Figure 3.**
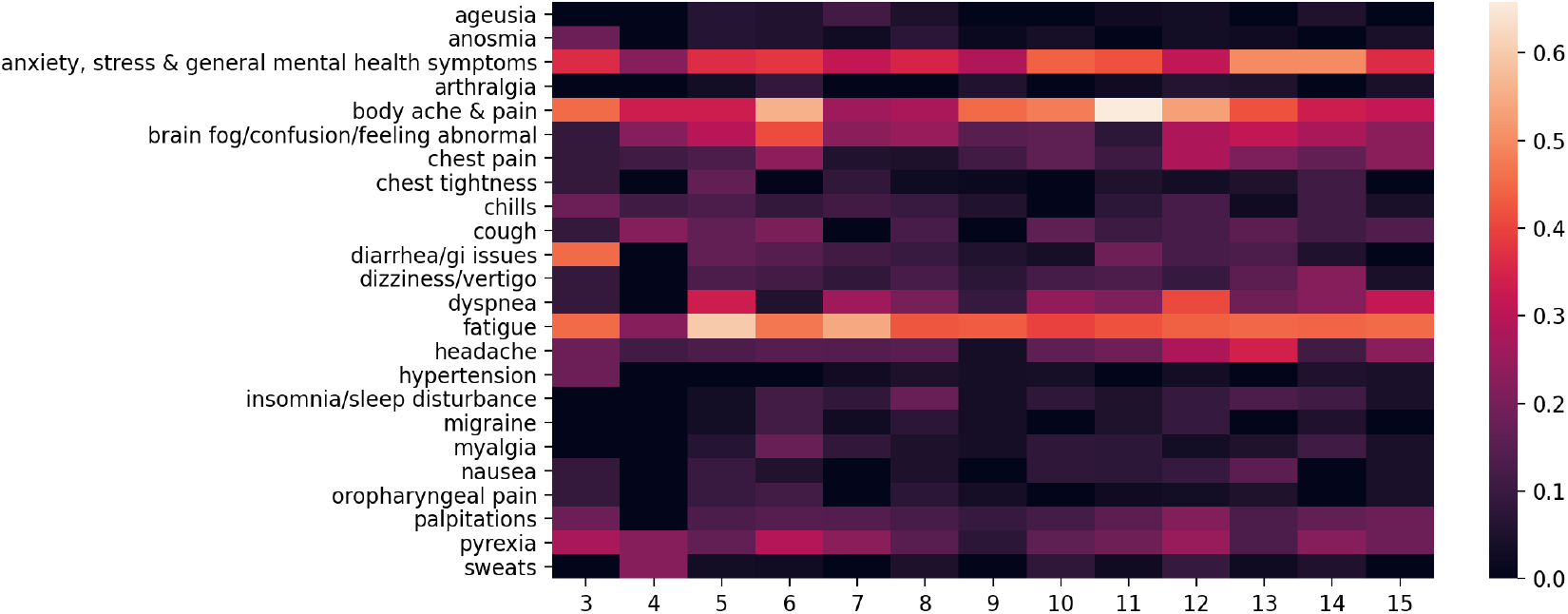
Heatmap of relative frequencies of symptoms from months 3 to 15.

## DISCUSSION AND CONCLUSIONS

While there is still limited research data available about long-COVID symptoms, our findings broadly agree with findings from recently-published studies. The ranges for the symptom frequency distributions in Table 1 are wide, which is likely due to the differing patient populations included in the studies. This also suggests that the long-COVID experiences of patients can vary considerably compared to acute COVID-19 infections, and further research is essential to better understand the chronic manifestation of this syndrome. Figure 3 shows that patients continue to experience certain symptoms, such as *fatigue, dyspnea, aches & pains*, and *mental health* symptoms up to 15 months following acute infection. Continuing surveillance of this cohort may reveal how the symptoms resolve, if at all. From these findings, it is evident that as some countries manage to lower new COVID-19 infections through vaccinations, an ongoing global challenge will be to address complications associated with long-COVID.

With limited scientific data available about long-COVID and with medical care globally primarily focusing on treating acute COVID-19 cases, many long-COVID sufferers are turning to social media to discuss their persistent symptoms, finding other sufferers with similar symptoms, and identifying potential solutions to their debilitating symptoms and improving quality of life. It is possible, and perhaps likely, that many patients will continue to suffer from long-COVID symptoms in the future, and social media will be an invaluable resource for obtaining early, crowdsourced insights about the topic. While manually reviewing many of the posts from our chosen subreddit, we found numerous posts where the users expressed their frustrations regarding their healthcare providers not understanding the extent of their sufferings and/or not being able to diagnose the underlying reasons behind their persistent symptoms. For example, many users who reported *dyspnea* stated that they had no issues with their lungs during the acute phases of their COVID-19 infections, and their care providers, who relied on standard imaging methods for diagnosis, dismissed their sufferings since imaging did not reveal anything wrong. We also found that many users explained that their symptoms were not continuous, but cyclic—disappearing and returning from time to time. Many users also discussed that their long-COVID symptoms were preventing them from getting back to their usual lives and work, adding to their mental health problems.

Our study has several notable limitations: the detection of symptoms is not 100% accurate, users may not report all the symptoms they experience (leading to underestimation of symptom prevalence), Reddit users tend to be younger compared to the general population, and rare symptoms may have been missed during our lexicon preparation. In our future work, we will attempt to analyze, from social media, how long-COVID typically progresses, the distribution of time spans at which users report the resolution of symptoms, and treatment regimens that appear to help. Social media data, including from Reddit and Twitter, may help the medical community to better understand long-COVID from the perspective of patients and thus enable them to improve the care they provide. Considering the large volume of data that is generated in social media about this topic, it is necessary to develop automated methods involving NLP for long-term surveillance. To aid future research, we have made the lexicon developed for this study publicly available.

## Data Availability

Lexicon associated with the article is publicly available.

https://sarkerlab.org/covid_sm_data_bundle

## Footnotes

† We grouped such symptoms into one because users often use non-standard expressions for these and it is difficult to separate such symptoms in a more fine-grained manner.

‡ The most commonly used phrase by the users is *brain fog*.

